# High Viral Specific Antibody Convalescent Plasma Effectively Neutralizes SARS-CoV-2 Variants of Concern

**DOI:** 10.1101/2022.03.01.22271662

**Authors:** Maggie Li, Evan J. Beck, Oliver Laeyendecker, Yolanda Eby, Aaron AR Tobian, Patrizio Caturegli, Camille Wouters, Gregory R. Chiklis, William Block, Robert McKie, Michael Joyner, Timothy D. Wiltshire, Allan B. Dietz, Thomas J. Gniadek, Arell Shapiro, Anusha Yarava, Karen Lane, Daniel Hanley, Evan M. Bloch, Shmuel Shoham, Edward R. Cachay, Barry R. Meisenberg, Moises A. Huaman, Yuriko Fukuta, Bela Patel, Sonya L. Heath, Adam C. Levine, James H. Paxton, Shweta Anjan, Jonathan M. Gerber, Kelly A. Gebo, Arturo Casadevall, Andrew Pekosz, David J. Sullivan, the CSSC group

## Abstract

The ongoing evolution of SARS-Co-V2 variants to omicron severely limits available effective monoclonal antibody therapies. Effective drugs are also supply limited. Covid-19 convalescent plasma (CCP) qualified for high antibody levels effectively reduces immunocompetent outpatient hospitalization. The FDA currently allows outpatient CCP for the immunosuppressed. Viral specific antibody levels in CCP can range ten-to hundred-fold between donors unlike the uniform viral specific monoclonal antibody dosing. Limited data are available on the efficacy of polyclonal CCP to neutralize variants. We examined 108 pre-delta/pre-omicron donor units obtained before March 2021, 20 post-delta COVID-19/post-vaccination units and one pre-delta/pre-omicron hyperimmunoglobulin preparation for variant specific virus (vaccine-related isolate (WA-1), delta and omicron) neutralization correlated to Euroimmun S1 IgG antibody levels. We observed a 2-to 4-fold and 20-to 40-fold drop in virus neutralization from SARS-CoV-2 WA-1 to delta or omicron, respectively. CCP antibody levels in the upper 10% of the 108 donations as well as 100% of the post-delta COVID-19/post-vaccination units and the hyperimmunoglobulin effectively neutralized all three variants. High-titer CCP neutralizes SARS-CoV-2 variants despite no previous donor exposure to the variants.

**Key points:** All of the post-delta COVID-19/post vaccination convalescent plasma effectively neutralizes the omicron and delta variants.

High-titer CCP and hyperimmunoglobulin neutralizes SARS-CoV-2 variants despite no previous donor exposure to the variants.

## Introduction

Commercial serologic assays predictive of SARS-CoV-2 and variant neutralization are important for effective clinical use of COVID-19 convalescent plasma (CCP), because substantial heterogeneity exists in CCP donor responses with higher antibody levels associated with virus neutralization.^1-3^ The SARS-CoV-2 omicron variant BA.1 rendered many monoclonals ineffective in laboratory virus neutralization tests, necessitating their removal as outpatient monoclonal therapies for acute COVID-19.^4,5^ While Sotrovimab retained activity against omicron BA.1, activity was lost against the BA.1.1 and BA.2 Omicron variants. Tixagevimab/cilgavimab (Evusheld), approved only for post-exposure prophylaxis, was ineffective at neutralizing omicron BA.2 and showed reduced activity towards omicrons BA.1 and BA.1.1.^6^ Bebtelovimab neutralizes omicron BA.1, but will have limited availability.^7^

A recent large clinical trial over the period from June 2020 to October 2021 demonstrated that early outpatient CCP reduced the risk of hospitalizations by more than half.^8^ Ninety percent of the trial CCP donor units were collected prior to January, 2021 representing pre-alpha/delta/omicron variants. In December 2021, the FDA extended CCP from hospital use to immunosuppressed outpatients, while simultaneously increasing commercial serologic benchmarks by 1.5-fold for CCP qualification.^9^

Recent omicron variant studies measured virus neutralization without concomitant commercial serologic testing for general antibody levels.^5^ Wang and colleagues measured a 10-fold virus neutralization reduction from SARS-CoV-2 wild type to omicron in 16 individual plasma samples obtained from January to March 2020 in China.^10^ Röllser *et al* examined only 10 CCP units from donors infected with variants and showed a lack of CCP neutralization of omicron but substantial neutralization with post COVID-19 post vaccination plasma.^11^ These studies have only tested a few samples and do not correlate the results to a commercial assay necessary to qualify therapeutic CCP units. Considering that CCP donors vary significantly in terms of virus neutralization capacity, the use of commercial assays that predict neutralization of SARS-CoV-2 and its variants becomes important to select the optimal therapeutic CCP units.

We examined a large group (over 100) of CCP units for their ability to neutralize three SARS-CoV-2 isolates/variants (WA-1, delta, and omicron), correlating results to the overall plasma levels of spike-S1 antibodies.

## Methods

### Participants

The study tested a total of 129 samples: 108 CCP donor units, 20 post-delta COVID-19/post-vaccination donor units and one pre-delta/pre-omicron hyperimmunoglobulin preparation. After the outpatient clinical research trial transfusions were complete, 108 remnant qualified (positive antibody presence after 1:320 dilution using a validated spike protein ELISA assay in a CLIA certified laboratory) donor plasma units were available for WA-1, delta, and omicron virus neutralizations.^8^ 94 (88%) pre-delta/pre-omicron units were donated before March 1, 2021; the remaining were collected in March 2021. Approvals were obtained from the Institutional Review Boards at Johns Hopkins University School of Medicine as single IRB for all participating sites and the Department of Defense (DoD) Human Research Protection Office. All participants provided written informed consent. The post-delta COVID-19/post-vaccination plasma aliquots were obtained from Innovative Transfusion Medicine after research informed consent. The collections occurred late August through October 2021 representing delta variant infection.

The hyperimmunoglobulin sample was prepared from 101 CCP units, collected between June 16, 2020 and January 13, 2021 (representing pre-delta/pre-omicron plasma) from recovered patients by an apheresis collection technique following Mayo Clinic Blood Donor Center’s standard operating procedures for apheresis plasma donations. Pooled plasma was isolated by fast protein liquid chromatography using protein A resin to capture IgG. Briefly, plasma was loaded on the protein A resin at a neutral pH, washed to remove low affinity proteins and eluted at low pH. Final product was formulated as a 5% protein solution in glycine/acetic acid.

#### Microneutralization studies

Plasma neutralizing antibodies (nAbs) were determined as described for SARS-CoV-2.^1,12^ WA-1 (SARS-CoV-2/USA-WA1/2020 EPI_ISL_404895) was obtained from BEI Resources while the, delta (hCoV19/USA/MD-HP05660/2021 EPI_ISL_2331507) and omicron (hCoV19/USA/MD-HP20874/2021 EPI_ISL_7160424) variants were isolated from COVID-19 patients at Johns Hopkins Hospital as previously described.^13^ The nAb titer was calculated as the highest serum dilution that eliminated the cytopathic effect in 50% of the wells (NT50) and the area under the curve (AUC) was calculated using Graphpad Prism. The total plasma levels of antibodies against spike region S1 were measured using the ELISA Euroimmun assay as described.^14^

## Results/Discussion

Compared to the WA-1 isolate, we observed a 2-to 4-fold decrease in virus neutralization of delta and 20-to 40-fold neutralization decrease of omicron in all plasma samples (108 pre-delta/pre-omicron CCP units, the 20 post-delta COVID-19, post vaccination units and pre-delta/pre-omicron hyperimmunoglobulin) (Fig. 1). The 108 research trial remnant units with Euroimmun AU over 3.5 have an 85% rate of positive virus neutralization for WA-1 and delta. However, Euroimmun over 10 AU was necessary to retain similar neutralization for omicron (Fig.1). All of the post-delta COVID-19/post-vaccination donor plasma as well as the hyperimmunoglobulin, with Euroimmun AU over 10 effectively neutralized the WA-1 isolate, delta and omicron variants. As a benchmark for clinical effectiveness, the early treatment CCP trial, successful at preventing hospitalizations principally with unvaccinated ancestral virus, had 251 (81%) of pre-delta/pre-omicron donor units with Euroimmun AU over 3.5 with a geomean average of 37 WA-1 neutralizing antibody IU/mL, while for all the transfused units the neutralizing antibody geomean was 26 IU/mL.^8^ The 108 CCP plasma units had an average of neutralization titer against WA-1 of 28 IU/mL. However, sorting to increasing Euroimmun AUs from greater than 3.5, to 7 and to 10 AUs, increased two-fold and ten-fold respectively WA-1 neutralization to 218 and 709 IU/mL (Fig 2). Again, we saw a similar drop in virus neutralization to delta and omicron variants when results were sorted by Euroimmun value. All of the post-delta COVID-19/post vaccination samples measured Euroimmun AU over 10, with WA-1 neutralization at geomean 1598 and still retained omicron neutralization, but with comparable reduction in activity of two-fold with delta and then 20-fold from the WA-1 isolate (Fig. 2). The pre-delta/pre-omicron hyperimmune globulin IgG showed a similar fold decrease in neutralization across the three isolates (Fig. 2). The wider span of both high and low levels of neutralization antibodies in CCP may account for the larger decrease in delta activity compared to the narrower high range of virus neutralizations with the post-delta COVID-19/post-vaccination units. This data suggests that Euroimmun AU over 5 would be effective for delta and over 10 for omicron CCP therapy.

**Figure 1.**
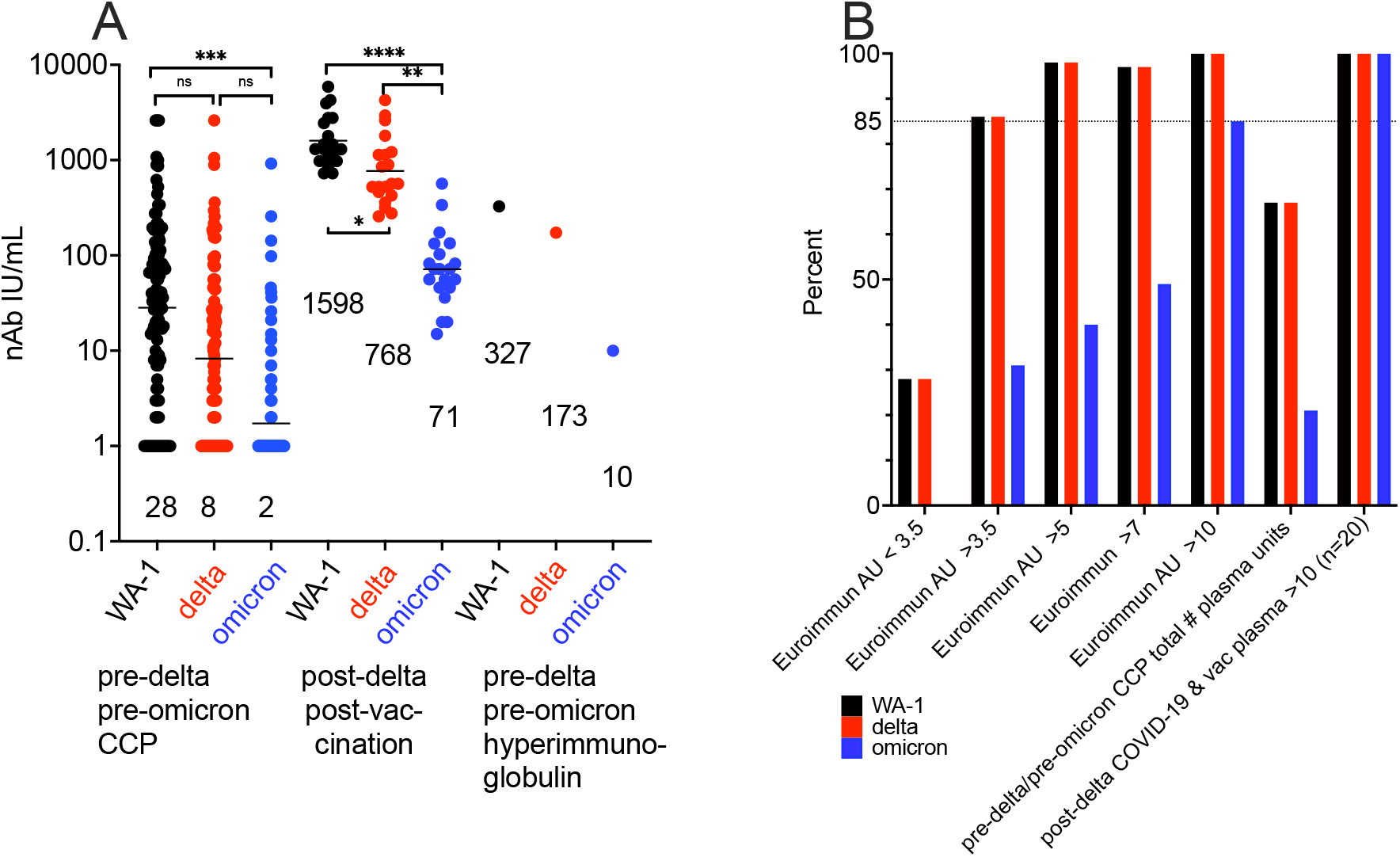
Reduction in virus neutralization sorted by plasma type as well as an increase by Euroimmun antibody levels. A) WA-1, delta and omicron virus microneutralization sorted by the 108 CCP units, post-delta COVID-19, post vaccination and hyperimmunoglobin. IU/mL geometric means are shown above x-axis. B) Antibody levels over 3.5 Euroimmun AU show 85% microneutralization with WA-1 and delta while omicron neutralization requires Euroimmun AU over 10 for 85% of the 108 CCP donors. Post-delta COVID-19/post vaccination retains 100% virus neutralization for the three variants. Virus neutralization is any positive IU/mL over 1. ****p<0.0001;***p<0.001; **p<0.01

**Figure 2.**
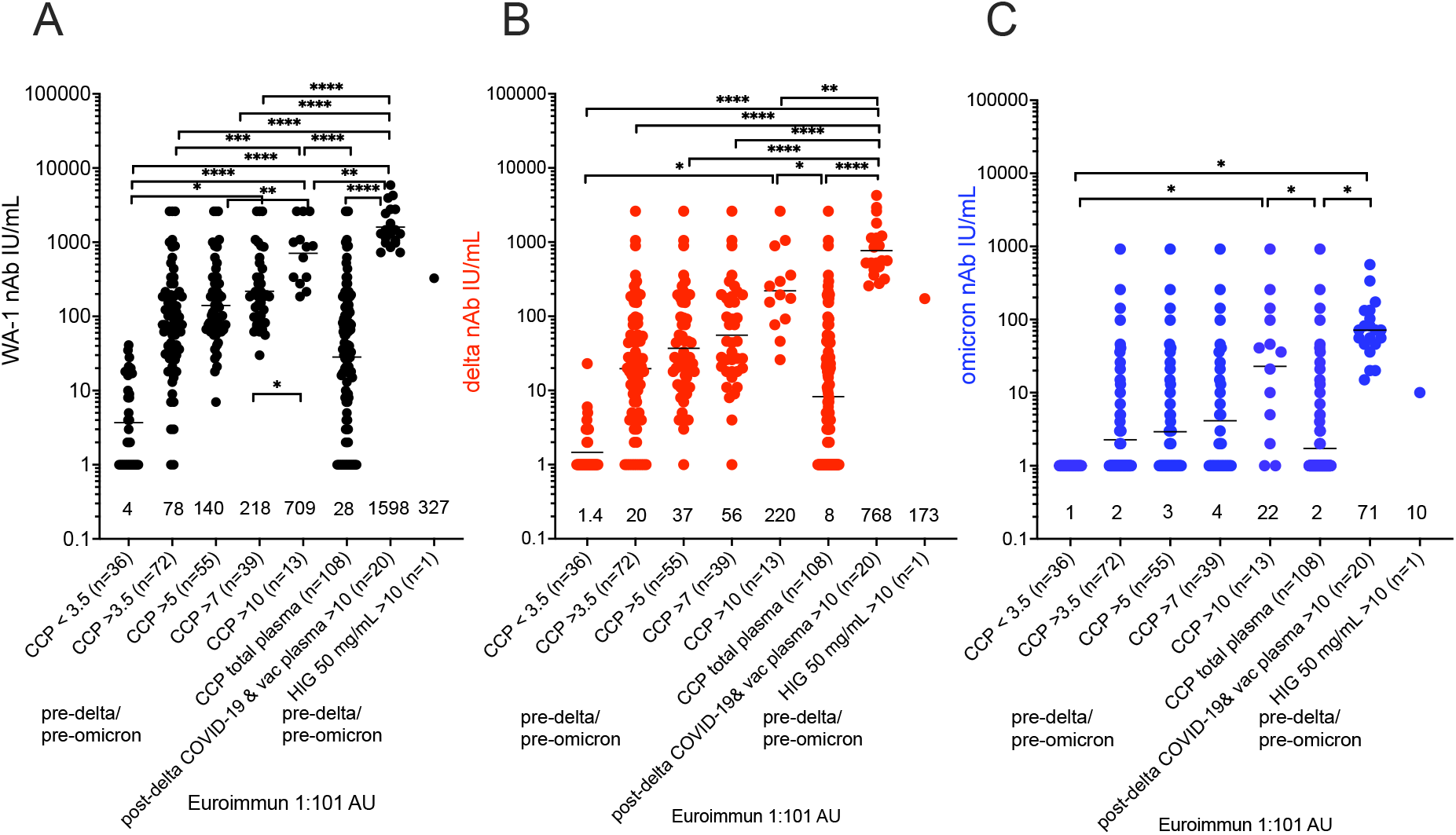
Sorting higher Euroimmun categories indicates virus neutralization to WA-1, delta and omicron. WA-1 (A), delta (B) and omicron (C) virus microneutralization measured in CCP, post-delta COVID-19/post-vaccination and hyperimmune globulin (HIG) sorted for viral-specific antibody levels by Euroimmun arbitrary units (AU) at 1:101 dilution. IU/mL geometric means are shown above x-axis. ****p<0.0001;***p<0.001; **p<0.01; *p<0.05

Commercial serologic assays were adjusted approximately 1.5-fold by the FDA to be in the effective range for omicron.^9^ The Euroimmun assay is one of many that can be utilized for donor plasma qualification. Many published studies on variant virus neutralization by CCP characterize 10 to 30 individual donors deeming them inactive against variants with small sample sizes. In contrast, here we compared over 100 CCP donor units to show that high viral specific antibodies defined by commercial serologic tests indicates omicron neutralization. The post-delta COVID-19/post-vaccination donor units and hyperimmune globulin retain broad activity against variants through omicron indicating its potential efficacy as a CCP treatment. This study did not characterize omicron convalescent plasma neutralization of older variants. We posit that high titer polyclonal CCP donors characterized with commercial serologic assays with a mismatch to existing variants still neutralizes SARS-CoV-2 variants as well as lower titer donor plasma units with a direct match of plasma to virus variants. These high titer units can be considered for CCP therapy in the face of variants.

## Data Availability

All data produced in the present study are available upon reasonable request to the authors

## Acknowledgments

We gratefully acknowledge the generous contributions of the study participants and plasma donors who gave of their time and specimens.

## Funding

The study was funded principally by the U.S. Department of Defense’s Joint Program Executive Office for Chemical, Biological, Radiological and Nuclear Defense (JPEO-CBRND), in collaboration with the Defense Health Agency (DHA) (contract number: W911QY2090012), with additional support from Bloomberg Philanthropies, State of Maryland, the National Institutes of Health (NIH) National Institute of Allergy and Infectious Diseases (NIAID) 3R01AI152078-01S1, NIH NIAID contract N7593021C00045 to the Johns Hopkins Center of Excellence in Influenza Research and Response (JH CEIRR), NIH National Center for Advancing Translational Sciences U24TR001609 and UL1TR003098, Division of Intramural Research NIAID NIH, Mental Wellness Foundation, Moriah Fund, Octapharma, HealthNetwork Foundation and the Shear Family Foundation.

## Authorship

Contribution: DS, AC, AP, TG & AT designed experiments; ML, CW, CB, AP, EB, YE performed experiments; MJ, TW and AD purified the hyperimmunoglobulin; GC, WB and RM provided post-delta COVID-19, post-vaccination plasma; AY, KL managed trial research blood bank; AS, EB, TG were contributing blood bankers; EC, BM, MH, YF, BP, SH, AL, JP, SA, JG contributed to the conduct of the related clinical trial, DS, KG, DH, EB, AT and SS organized clinical trial studies as source of CCP; DS, ML, AP analyzed data and wrote the manuscript with input from all authors and all authors approved the final version of the manuscript. The full COVID-19 Serologic Studies Consortium (CSSC) authors are listed in the appendix.

## Conflict-of-interest disclosure

TG-paid consultant for Fresenius Kabi; GC and RM are on the Board of Innovative Transfusion Medicine; WB-Board of Blood Centers of America; AC-Scientific Advisory Board of Sabtherapeutics (cow-derived human immunoglobulins COVID-19 treatment and other infectious diseases) and Ortho Diagnostics Speakers Bureau; EB-member of the FDA Blood Products Advisory Committee. All other authors report no relevant disclosures.

## Appendix

The members of the COVID-10 Serologic Studies Consortium (CSSC) are: Anne Arundel Medical Center (Barry R. Meisenberg); Ascada Research (Matthew Abinante, Kevin Oei); Baylor College of Medicine (Yuriko Fukuta); MedStar Georgetown University Hospital (Seble Kessaye); Johns Hopkins Center for American Indian Health (Laura L. Hammitt, Catherine G. Sutcliffe) Johns Hopkins Bloomberg School of Public Health (David J Sullivan, Bryan Lau, Atika Singh, David M. Shade, Stephan Ehrhardt, Sheriza N. Baksh, Andrew Pekosz, Sabra L. Klein, Arturo Casadevall); Johns Hopkins University (Kelly A Gebo, Shmuel Shoham, Evan M. Bloch, Christi E. Marshall, Anusha Yarava, Karen Lane, Nichol A. McBee, Amy L. Gawad, Nicky Karlen, Daniel E. Ford, Douglas A. Jabs, Lawrence J. Appel, Oliver Laeyendecker, Aaron A.R. Tobian, Daniel F. Hanley); Lifespan/Brown University Rhode Island Hospital (Adam C. Levine); Mayo Clinic, Phoenix (Janis E. Blair); MedStar Washington Hospital Center (Aarthi Shenoy); NorthShore University HealthSystem (Giselle S. Mosnaim, Thomas J. Gniadek); The Bliss Group (Michael Roth); The Next Practice Group (Colin Foster); University of California Los Angeles (Judith S. Currier); University of Alabama at Birmingham (Sonya L. Health); University of California, Irvine Health (Donald N. Forthal); University of California, San Diego (Edward R. Cachay, Elizabeth S. Allen); University of Cincinnati Medical Center (Moises A. Huaman); University of Massachusetts Worcester (Jonathan M. Gerber); University of Miami (Shweta Anjan); University of New Mexico (Jay S. Raval); University of Rochester (Martin S. Zand); University of Texas Health Science Center at Houston (Bela Patel); University of Utah Health (Emily S. Spivak, J. Robinson Singleton); Vassar Brothers Medical Center (Valerie C. Cluzet, Daniel Cruser); Wayne State University (James H. Paxton); Western Connecticut Health Network, Danbury Hospital (Joann R. Petrini, Patrick B. Broderick, William Rausch, MarieElena Cordisco); Western Connecticut Health Network, Norwalk Hospital (Jean Hammel, Benjamin Greenblatt)

## Notes

### Funding Statement

The study was funded principally by the U.S.Department of Defense Joint Program Executive Office for Chemical, Biological, Radiological and Nuclear Defense (JPEO-CBRND), in collaboration with the Defense Health Agency (DHA) (contract number: W911QY2090012), with additional support from Bloomberg Philanthropies, State of Maryland, the National Institutes of Health (NIH) National Institute of Allergy and Infectious Diseases (NIAID) 3R01AI152078-01S1, NIH NIAID contract N7593021C00045 to the Johns Hopkins Center of Excellence in Influenza Research and Response (JH CEIRR), NIH National Center for Advancing Translational Sciences U24TR001609 and UL1TR003098, Division of Intramural Research NIAID NIH, Mental Wellness Foundation, Moriah Fund, Octapharma, HealthNetwork Foundation and the Shear Family Foundation. The study sponsors did not contribute to the study design, the collection, analysis, and interpretation of data, and the decision to submit the paper for publication.

### Author Declarations

Approvals were obtained from the Institutional Review Boards at Johns Hopkins University School of Medicine as single IRB for all participating sites and the Department of Defense (DoD) Human Research Protection Office. All participants provided written informed consent. The post-delta COVID-19/post-vaccination plasma aliquots were obtained from Innovative Transfusion Medicine after research informed consent.

